# Supervised and self-directed technology-based dual-task exercise training programme for older adults at risk of falling – Protocol for a feasibility study

**DOI:** 10.1101/2024.11.19.24317600

**Authors:** Prerna Mathur, Helen Thomas, Angela Cooper, Magdalena Chechlacz, Afroditi Stathi, Victoria Goodyear, Caroline Miller, Taylor Krauss, Natalie Ives, Laura Magill, Philip Kinghorn, Daisy Wilson, Shin-Yi Chiou

## Abstract

Falls among older adults pose a significant public health challenge, as they lead to severe outcomes such as fractures and loss of independence. Research has shown that training cognitive function and balance simultaneously, termed Dual-Task (DT) training, improves mobility and reduces fall risks in older adults. This study aims to evaluate the feasibility and acceptability of a blended supervised and self-directed technology-based DT training programme for older adults who have high risk of falling. This is a single-arm, non-randomised feasibility study employing quantitative and qualitative methods. Fifty healthy adults aged 65 years or above will be recruited from the NHS primary and secondary care pathways and from the community. Participants will undergo supervised cognitive and balance DT training for 12 weeks, followed by self-directed DT training for an additional 12 weeks. The cognitive training will be delivered using a commercial mobile application (app) available from the AppStore or Google Play. The balance training will involve static (Marching on the spot, Tandem Stand, Hip Abduction & Extension, Squats, Tiptoe Stand, and Pendulum/Sideways Sway) and dynamic (Figure of Eight Walk, Walking Forwards and Backwards, Lunges, Functional Reach, Toe Tapping, Upper Limb Strength Exercises, and Side-Steps/Simple Grapevine) exercises focused on improving balance, postural stability and strength. Feasibility outcomes will be recruitment, adherence, usage of the app, and attrition. Outcomes measure data, that will be collected at baseline and at 24 weeks, includes the Timed-Up and Go (TUG) test (likely primary outcome in any future trial), along with self-reported questionnaires assessing cognition, fear of falling, quality of life, healthcare service usage, and the self-reported number of falls. Focus group interviews will be conducted with thirty participants and thirty healthcare professionals for in-depth exploration of the feasibility and acceptability of the DT training programme.

**Trial registration number:** ISRCTN15123197

## Introduction

Falls among older adults represent a significant public health issue. Around 1 in 3 adults aged over 65, and half of adults aged over 80, have at least one fall a year, and in half of such cases the falls are recurrent (1, 2). Falls can lead to severe consequences such as hip fractures, which often result in reduced mobility and loss of independence (3). An estimated £4.4 billion is spent annually on treating fractures and fall-related injuries (4). The prevalence of falls and associated complications necessitates the development and implementation of effective intervention strategies aimed at prevention.

Evidence suggests that age-related declines in cognition and mobility are risk factors for falls in older adults(5–7). Studies reveal that older adults with better mobility perform better in assessments of global cognition, executive function, memory and processing speed (7–11), suggesting the interplay between cognition and mobility. Research has shown that the decreasing ability to multitask is associated with increasing risk of falling in older adults (12, 13) and that training cognitive and physical function simultaneously, termed Dual-Task (DT) training, is superior to single-task (i.e., only physical function) training or no training for improving walking speed in older adults (1, 14).

The cognitive element of DT training can be delivered via technology (e.g., mobile apps), enabling professionals to select cognitive exercises that are suitable to be combined with physical exercises, allowing for the DT training to be self-directed and performed outside of clinical environments. Mobile apps are interactive, provide instant feedback, and can send reminders to users. These features promote engagement and adherence to exercise(15). Previous studies reported improved balance and walking speed after self-directed, home-based DT training programmes with mobile apps in older adults, with excellent adherence rate (85-90%) (16, 17). These findings support the notion that technology-based exercise (not limited to DT exercise) may be a sustainable means of promoting physical activity and preventing falls in older adults(18–20).

Whilst a purely technology and home-based exercise programme seems attractive, it may appear daunting to some older adults, such as those who are reported to have lower levels of knowledge and competence in using mobile apps(21, 22). Barriers to the use of technology in the medical context amongst older adults have been reported(23). A blended approach, combining supervised sessions with self-directed sessions, can help mitigate the challenges older adults might face while using the mobile app by providing initial hands-on guidance and ongoing support. The gradual transition from supervised to self-directed exercises allows participants to build confidence and familiarity with the technology, ultimately improving their engagement and adherence(24).

While it is evident that a DT programme can be administered well under supervision(14, 25), we have limited knowledge on how to best assure its quality, outcomes, and compliance when administered unsupervised with the support of technology, such as a mobile app. Therefore, the primary aims of this study are to evaluate; (1) the acceptability of a technology-based DT programme with blended supervised and self-directed approaches in older adults at higher risk of falling and (2) to determine the feasibility of the programme to be adopted by the National Healthcare System as a treatment for falls prevention and management in older adults in the United Kingdom.

### Objectives

1. To evaluate whether a blended supervised and self-directed DT training programme delivered via a mobile app is acceptable to older adults living in the community who have had recurrent falls in the past 12 months.
2. To assess the feasibility of the technology-based blended DT training to be implemented in the NHS for falls prevention and management in older adults.

The information collected will inform the design of a larger scale Randomised Controlled Trial to evaluate the clinical and cost-effectiveness of the blended intervention in older adults aged 65 years and above at risk of falls.

## Methods and analysis

### Study Design

This is a single-arm, non-randomised feasibility study of a technology-based, blended DT training programme in older adults with a history of falls (Fig 1) outlines the schedule of enrolment, interventions and assessment and provides insight into the trial process and data collection. The flow of the study is shown in Fig 2. This study was approved by the East of England - Cambridge East Research Ethics Committee (REC reference: 24/EE/0059) and registered on the International Standard Randomised Controlled Trials Number Registry (ISRCTN15123197) on April 16, 2024. The study protocol is reported following the SPIRIT check list (S1 Appendix).

**Fig 1.**
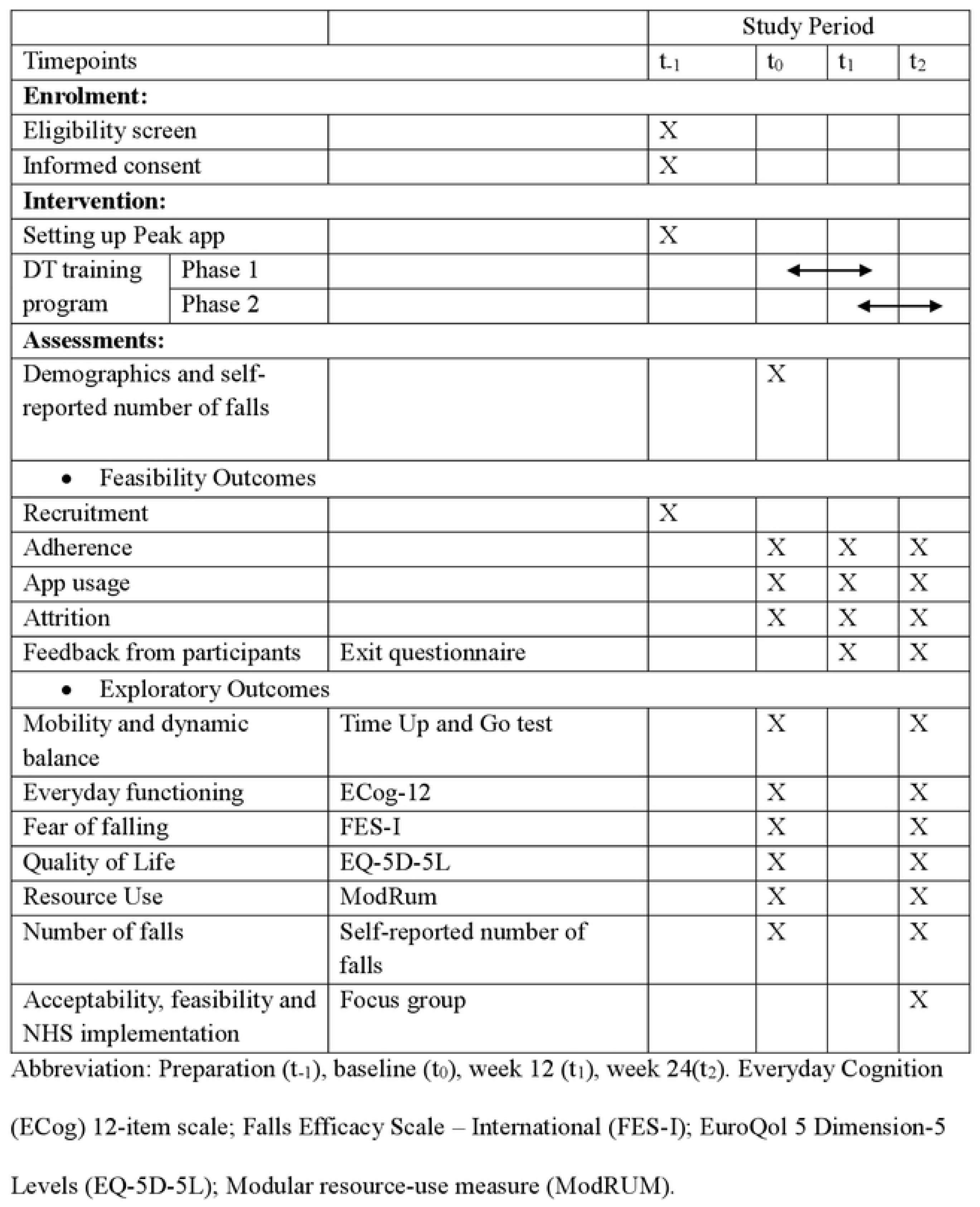
Schedule of enrolment, interventions and assessment.

**Fig 2.**
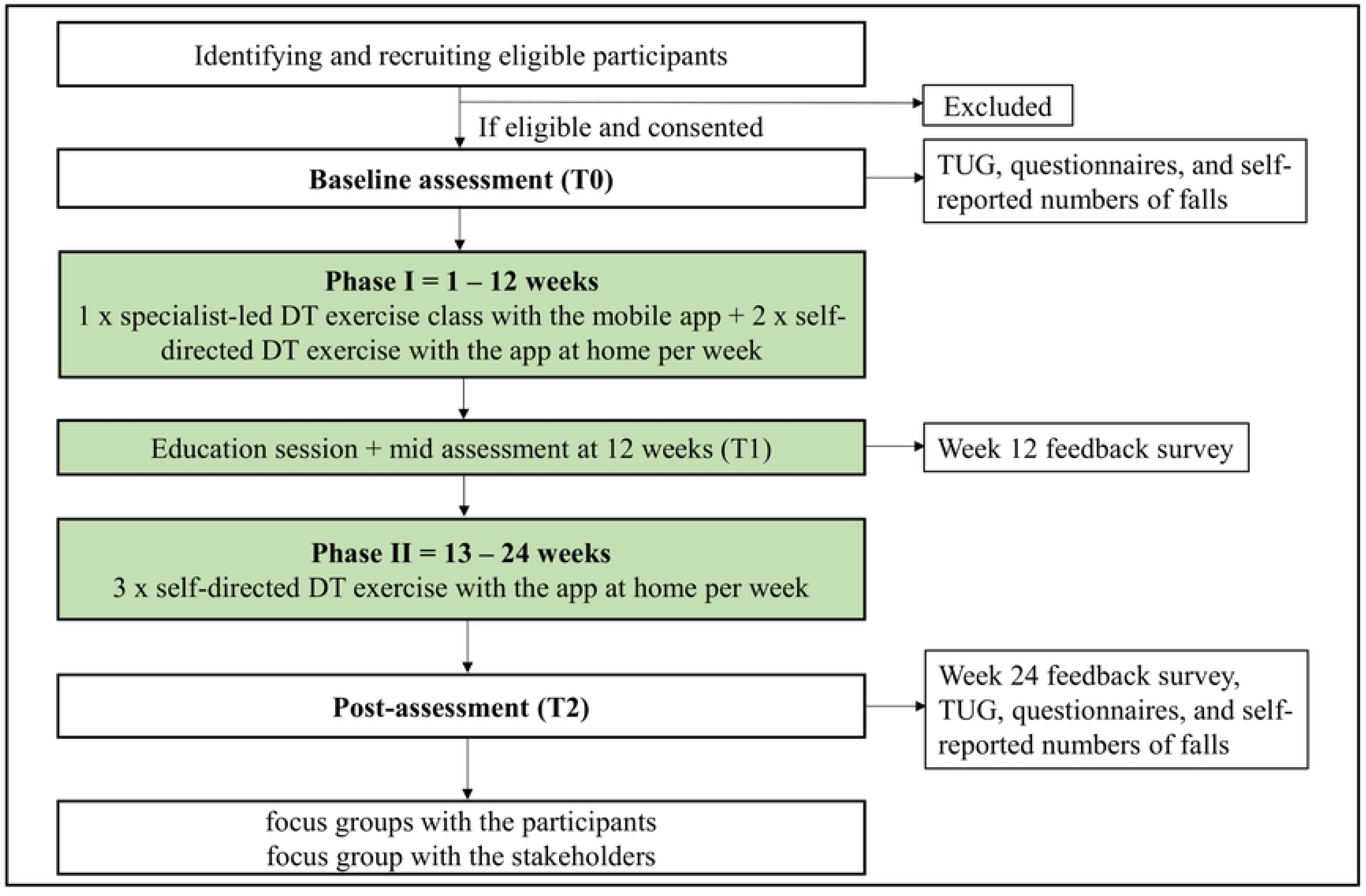
Study Flow Diagram. DT: dual-task; TUG: Timed Up & Go.

### Setting

The study will be conducted in a community setting in Birmingham, UK. There are two phases: phase 1 is supervised cognitive and balance DT training for 12 weeks at local community centres, followed by a phase 2 which is self-directed DT training for an additional 12 weeks.

### Participants and recruitment

Participants will be recruited from primary and secondary care pathways in the National Health Services (NHS) in England and from the community in the West Midlands. The study will be promoted via a number of methods including general practitioners sending text messages to patients, healthcare professionals identifying eligible patients in the falls clinics, and study flyers displayed in relevant organisations, such as retirement villages, charity organisations, and interest groups. Interested individuals from the NHS and from the community will be directed to an online participant information sheet (PIS) or given a copy of the PIS by their healthcare professional and by the research team, respectively. They can contact the research team to request further information of regarding participation and/or to ask any study-related questions they may have. If they are interested and willing into be enrolled in the study, the research team will confirm the eligibility of the potential participant via telephone or a face-to-face appointment, explain the study and what is involved, should they agree to take part, answer any questions they have. Written informed consent (S2 Appendix) will be obtained from all participants at community venues where the baseline assessment and supervised intervention will be held. Recruitment was started on 29/04/2024 and is ongoing.

#### Eligibility Criteria

The inclusion criteria are:

1) aged 65 years and above
2) able to give informed written consent
3) having sufficient cognition/hearing/vision to follow instructions of the assessment and the exercise programme
4) able to stand with one hand support on the current walking aid for at least 60 seconds
5) able to stand up from a chair independently and walk for 6 meters independently with the current walking aid
6) able to use toilet independently
7) having access to a smartphone or tablet compatible to the Peak-brain training app(26, 27) on iOS or Android devices
8) had two or more falls in the last 12 months.

The exclusion criteria are:

1) having an unstable or acute medical condition (such as fractures, acute coronary syndromes) that preclude exercise participation
2) suffer from a progressive neurological condition (such as Parkinson’s disease or multiple sclerosis)
3) not recommended to undertake any forms of exercise by their doctors
4) currently participating in a different research study for managing their fall risks.

#### Sample size

The sample size (n=50) was chosen based on published literature where a sample size of between 50 and 100 participants (total; i.e. 25 to 50 per group) is recommended for pilot studies(28–30). If we identify 100 eligible participants, we will be able to estimate a participation rate of 50% within a 95% confidence interval of +/−9.8%. With a sample size of 50 participants, we will be able to estimate an attrition rate of 20% within a 95% confidence interval of +/−11%.

### Intervention

All participants will receive the DT training programme for 24 weeks. The programme has two phases (Table 1). Phase 1 is 12 weeks long. It consists of supervised group exercises once a week, along with two other home-based sessions of self-directed, independent exercise similar to the class exercises. The group exercise class of 5-10 participants will be led by a physiotherapist 1 day/week. The classes will be approximately 50 minutes in duration and include 40 minutes of DT training with 10 minutes of warm-up and cool-down. The exercise classes will be held in local community centres that will be easy access for the participants.

**Table 1.** A summary of dual-task training programme.

| **Supervised group exercise** | **Self-directed, home-based exercise** |
| --- | --- |
| **Duration:** 50 minutes | **Duration:** 50 minutes |
| **Location:** local leisure/wellbeing centres | **Location:** at home |
| **Frequency:** one class/week (Weeks 1-12)<br>**Content:** 5 minutes warm-up, 40 minutes of DT training, 5 minutes cool down.<br>**DT training:**<br>• Cognitive exercises delivered via the Peak app.<br>• Strength and balance exercises delivered by a physiotherapist | **Frequency:** twice/week (Weeks 1-12); thrice/week (Weeks 12-24)<br>**Content:** 5 minutes warm-up, 40 minutes of DT training, 5 minutes cool down.<br>**DT training:**<br>• Cognitive exercises delivered via the Peak app.<br>• Strength and balance exercises in an exercise booklet |

Phase 2 is self-directed, with participants independently exercising at home using the app, 50 minutes per day, 3 days per week, for 12 weeks. Participants will have access to the Peak app(26) and will repeat the same exercises that they have performed in the phase 1but without supervision. This phase reinforces the idea of performing DT training as part of one’s routine for maintaining balance and mobility, thereby reducing the risk of recurrent falls.

#### DT training programme

The DT training programme requires participants to perform both cognitive tasks and balance and strength exercises simultaneously. The cognitive exercises are delivered via the Peak app(26) which is a commercial app that can be downloaded from the App Store (iOS) or Google Play (Android). The research team will assist participants to install the app on their own devices prior to the start of the first class. The PEAK app(26) has multiple games, and the choices of the games are based on two criteria: 1) whether a game can be safely performed with a balance exercise, and 2) the type of cognitive function being trained. For the second criterion, we prioritise the games that require executive function, memory, attention, and processing speed as they have been shown to relate to balance control and falls(31–34). Eighteen games covering the above cognitive domains were chosen within 4 categories of games available from the Peak app, Focus, Memory, Mental Agility, and Problem Solving (Must Sort, Rush Back, unique, Decoder; Perilous Path, Memory Sweep, Spin Cycle, Apprentice Wizard, Partial Match; True Color, Face Switch, Refocus, Turtle Traffic; Puzzle Box, Slider, Low Pop, Castle Block, Earth Defence).

For the physical exercise, the standardised static and dynamic balance and strength exercises routinely prescribed to older adults for falls management and prevention are included in the programme. The static exercises (Marching on the spot, Tandem Stand, Hip Abduction & Extension, Squats, Tiptoe Stand, and Pendulum/Sideways Sway; S3 Appendix) will be performed concurrently with the cognitive exercises (S4 Appendix). The dynamic exercises (Figure of Eight Walk, Walking Forwards and Backwards, Lunges, Functional Reach, Toe Tapping, Upper Limb Strength Exercises using a resistance band, and Side-Steps/Simple Grapevine) will be performed independently (i.e. not concurrently with ethe cognitive exercises) (S3 Appendix). These exercises were selected by a specialist physiotherapist from the University Hospitals Birmingham, considering the safety of participants in light of the dual task nature of the training programme.

#### Supervised group exercise classes

In the Phase 1 of the training programme, participants will be asked to attend an exercise class (of 5-10 participants) once a week for 12 weeks. In each class participants will be shown three static and three dynamic exercises respectively by a physiotherapist. The class will consist of a 5-minute warm-up at the start, after which participants will perform the three static exercises, followed by the three dynamic exercises, and finish with a 5-minute cool-down to complete the class. Each exercise will last for approximately 6 minutes, with short breaks between each exercise. The three static exercises will be performed with three different games from the Peak app for cognitive brain training.

The participant’s device will be placed on a height-adjustable music stand in front of the participant at an appropriate distance that allows them to use the app comfortably. There will be a chair placed next to the participant for support during the exercises if needed. The physiotherapist leading the class will provide individual support and adjustment to the exercises to each participant to ensure that everyone is able to engage with the DT exercise training as per their capacity. The Peak app adapts the games (increase/decrease of the difficulty level) based on the participant’s performance to maintain cognitive challenges as they progress. At the end of the class, participants can note down the exercises and variations performed in the class in an exercise diary.

#### Self-directed, home-based exercise

Participants will be instructed to perform the same DT exercises at home on another two days in the same week after each class in phase 1, and three times a week during phase 2, the self-directed, home-bases DT part of the programme. The scope of the blended approach is to allow participants time to learn the exercises and the technology, to develop an exercise routine, to improve their ability(35, 36), and potentially enhance their confidence to use the app. All participants will be given a handbook which has images and instructions of all the physical and cognitive exercises (S3 & S4 Appendices) to help them exercise at home. They will also have access to online videos demonstrating how to perform each exercise correctly and safely. Equipment required (height-adjustable music stand, and resistance bands) for exercising at home will be provided to the participants. Participants will be asked to record their adherence to the programme. In phase 1, participants will document, in an exercise diary provided to them at the start of the training programme, the cognitive games performed with the physical exercises. In phase 2, participants will be asked to document both the cognitive games and the physical exercises performed in the exercise diary. Participants will be able to contact the research team (via email or SMS) for technology assistance and/or to ask questions during the self-directed home-based exercise.

#### Educational session

An in-person educational session for falls awareness will be presented to the participants at the end of phase 1. This session will be conducted by the physiotherapist delivering the training programme. In the education session, participants will be reminded of the content of phase 2, access to the resources (i.e., the handbook and videos), and support they can receive if they run into any technical difficulties with the app.

Participants will also be asked to consider ways that may help them stay motivated in doing the exercise in the next 12 weeks, such as creating WhatsApp groups or using Facebook Messenger, and supplemented by face-to-face “coffee shop” get-togethers to stay in touch with other participants in the same age and/or living in the same area.

### Measures

#### Feasibility Outcome Measures

The feasibility outcomes of the study are whether the study is appealing to participants (assessed by the recruitment and retention rate) and if the intervention is acceptable (measured by adherence and usage of the app).

1. *Recruitment*. Recruitment will be calculated as (number recruited/number approached) × 100. The source of recruitment (e.g., primary care, secondary care, and community) will be documented to evaluate which recruitment route is the most appropriate for the main trial.
2. *Adherence.* Adherence will be calculated as: (self-reported number of exercise days/number of prescribed days) × 100 using the information recorded on the exercise diary.
3. *Usage of the app*. This will be analysed based on the self-reported number of games performed and the repetition of each game during the DT programme.
4. *Study attrition.* Attrition will be calculated as: [(number of dropouts)/ number recruited) × 100].
5. A self-reported online EXIT survey (S5 Appendix) will be completed by participants to provide feedback on the usability, perceived effectiveness and satisfaction from the training programme based on the feasibility assessment framework(37).

#### Stop-Go criteria

A ‘traffic light’ system based on recruitment, adherence, app usage and retention will be used to provide guidance as to whether the project should progress to a full trial with an internal pilot (green), progress to full trial with internal pilot with adaption (amber), or no progression (red).

- **Green:** recruitment ≥50%; adherence and usage of the app over the programme is ≥80% of the time (i.e., completed the physical exercises and cognitive exercises in 58 sessions or more out of a total of 72 sessions over 24 weeks); and retention is ≥80%. Focus group results support the feasibility and acceptability of the study and the blended intervention, and there are no safety concerns.
- **Amber:** recruitment 30-49%; adherence and usage of the app is 50-79%; and retention rate is 50-79% of the time. Focus group results indicate that changes in the design and delivery of the programme are required, and there are no safety concerns. Potential changes could be technical support required for the phase 2 where participants undertake the exercise independently.
- **Red:** recruitment <30%; adherence and usage of the app is <50%; and retention is <50% of the time. Focus group results do not support the study progressing to a larger-scale RCT, and/or there are safety concerns, which cannot be addressed.

Data from the feasibility study will directly feed into the above stop-go to provide guidance on progression to the full trial. Any decision will also be informed by the findings of the focus groups (see below qualitative methods for details).

#### Exploratory Outcomes

The study has three assessment periods – baseline (t_0_), Week 12 (t_1_), Week 24 (t_2_) where data collection is undertaken (Fig 2). Information on participant demographics (age, sex, ethnicity, post code and level of education) will be collected in order to describe the study participants and assess inclusion and reach of the study.

Data will be collected on the following outcomes which are planned outcome measures in any future trial in order to assess data collection procedures, data completeness and to help inform sample size calculation for the future trial.

1. *Mobility and dynamic balance.* Timed Up and Go (TUG)(38) with and without concurrent undertaking of a cognitive task will be carried out in the presence of the physiotherapist to evaluate mobility and dynamic balance of the participant. Participants will be asked to rise from a standard armchair, walk to a marker 3 meters away, turn, walk back, and sit down again. This will be completed three times, with the first TUG will be a practice round(39). For the TUG with cognitive tasks (TUG Cognitive)(38), participants will be asked to count backwards in sevens from a random start point while completing the TUG. The performance from the TUG and TUG Cognitive will be recorded.

The following self-reported questionnaires will be collected via an online survey (using REDCap software) at baseline and after the delivery of the training programme.

2. *Decline of cognition and everyday functional abilities.* Everyday Cognition scales short version (ECog-12; 12 questions in total)(40, 41) will be used to evaluate the decline of cognition and everyday functional abilities linking to independence in the activities of daily living. The ECog-12 questionnaire has 12 items on a scale from 1-4, being 1 (better or no change compared to 10 years ago) and 4 (consistently much worse). The results from this questionnaire will reflect any effect of the training programme on the cognitive function affecting daily living which declines with ageing.
3. *Self-efficacy in performing daily activities*. Falls Efficacy Scale-International (FES-I), having 16 items on a scale from 1-4, being 1 (not concerned at all), and 4 (very concerned), will be used to evaluate the confidence in the performance of activities of daily living in levels of fall risk (>24 points)(42).
3. *Quality of life.* The EQ-5D-5L will be used to assess the quality of life(43). The questionnaire has 5 items each with five response options, being 1 (no problems) and 5 (unable).
5. *Resource Use.* The ModRum questionnaire will be used to evaluate the use of healthcare services, providing basic health economics information of the intervention(44).
6. *Number of falls.* Self-reported number of falls.

#### Qualitative Methods

1. *Focus groups.* We will conduct face-to-face semi-structured focus groups with a purposive sample of 30 participants who have completed the programme to discuss the content and delivery of the training programme. We will also conduct online semi-structured focus groups with 30 healthcare professionals including GPs, district nurses, paramedics, and physiotherapists who are part of the NHS falls prevention care pathways in the West Midlands, via MS Teams. The healthcare professionals and physiotherapists involved in the study will be invited to take part in the focus groups to discuss the feasibility and deliverability of the training programme by the NHS. Discussions will be downloaded from MS Teams and transcribed verbatim. The focus group recordings will be coded using Nvivo9 software. Analysis will be deductive, informed by the study objectives, and will follow a thematic analysis approach. A deliberative approach will be used to interpret emerging themes with the diverse interdisciplinary author team, and this serves as a marker of quality (45, 46).

#### Statistical analysis

Given the study is a feasibility study, the analysis undertaken will mainly be descriptive. The feasibility outcome measures will be reported as proportions and percentages with 95% confidence intervals calculated. Outcome data will be summarised at baseline and follow-up using appropriate summary statistics. Exploratory analysis may compare the data form the two assessment time points using a paired t-test (depending on the distribution of the data) to provide preliminary data on the effect of the blended intervention. Analyses will be undertaken in SPSS(47).

#### Data Management

Paper based study records will be kept in locked cabinets within a locked office at the University of Birmingham. Electronic records will be stored on RedCap which is a secure data management software application managed by the Birmingham Centre for Observational and Prospective Studies (BiCOPS) at the University of Birmingham. The recordings from the focus groups will be held on a secure device and will be uploaded to the University of Birmingham server. Access to the files will be restricted and password protected.

#### Study management and safety

The DT training programme and the behavioural assessments are considered low risk. The research team will meet monthly to monitor the progress of the study, supervise the study, and discuss data and adverse events to ensure that the study is conducted in accordance with the approved protocol and regulations. Three members of the public will be recruited to the PPIE steering group and meet three times a year. The chief investigator will report the study progression to the PPIE steering group.

The DT programme was designed by experienced physiotherapists and a member of the public with lived experience to reduce risks and burdens as much as possible. Risks will be outlined in the PIS and verbally explained to all participants before they provide written informed consent. Participants will also be verbally reminded after each class on how to safely undertake the intervention at home.

#### Ethics and Dissemination

This study was approved by the Health Research Authority and Health and Care Research Wales (24/EE/0059). We anticipate that the study will be completed by 31/12/2025 and that results will be disseminated at international conferences and in published peer-reviewed journals in 2026.

## Discussion

Falls among older adults are a significant public health concern as they not only result in physical injuries but can also lead to psychological consequences, such as fear of falling, which further exacerbates the risk of subsequent falls(48). Given the multifactorial nature of falls, interventions that target both physical and cognitive function are crucial for fall prevention in older adults(49). This study seeks to evaluate the feasibility and acceptability of a technology-based dual tasking exercise training program designed specifically for older adults at risk of falling.

The information in this study will inform the feasibility and design of a larger scale randomised controlled trial evaluating the clinical and cost-effectiveness of the blended DT intervention in older adults at risk of falls. The knowledge generated will also have wider implications on other clinical conditions, where home exercise is a commonly used tool for maintaining/improving health. This study is especially relevant, given population ageing and the increased use of technology and mobile apps in older adults during the global pandemics.

However, there is limited understanding on how technology can facilitate dual tasking exercise programs for older adults. Data published by the Office for National Statistics show that 69% of adults over 65 years and 55% of adults over 75 years owned a smartphone in 2021(50). The number is likely to increase, meaning that it is now the time to evaluate how to best incorporate these digital tools into standard care and to determine if the quality, outcomes and compliance of technology-based interventions are comparable or superior to standard care.

## Data Availability

Data cannot be shared publicly because of confidentiality. Data are available by request from the corresponding author for researchers who meet the criteria for access to confidential data.

## Acknowledgement

We thank Liz Hensel and Tom Tierney from the PPIE steering group for their contributions in reviewing participant facing documents and feedback on the study design and management. We also thank Terry Hughes for setting up the RedCap database, which is critical to the successful management of this study.

## Contributors

Authors’ contributions SYC leads the study as the chief contributor. SYC, MC, AS, VG, CM, NI, LM, PK, and DW obtained the funding for the research. SYC, MC, AS, VG, CM, NI, LM, PK, DW, PM, AC, HT and TK contributed towards the design of the study. PM leads the coordination of the study as the trial manager. PM and TK are involved in collection of data. PM, SYC, NI, and PK are involved in the processing of data. NI will oversee and support the data and (any) statistical analysis. AS, VG, and PM are responsible for the qualitative analysis. SYC, CM and PM will manage the budget of the grant. PM and SYC drafted the manuscript with critical input from all other authors. All authors read and approved the final draft. All authors have consented to publication.

## Supporting information

**S1. Appendix. SPIRIT Checklist 2013**

**S2 Appendix. Consent Form**

**S3 Appendix. Exercise Handbook**

**S4 Appendix. Peak app Handbook**

**S5 Appendix. Exit questionnaire**

## References

1. Schoene D, Valenzuela T, Lord SR, de Bruin ED. The effect of interactive cognitive-motor training in reducing fall risk in older people: a systematic review. BMC Geriatr. 2014;14:107.

2. Falls – Overview2021 23-08-2024. Available from: https://www.nhs.uk/conditions/falls/#:∼:text=Around%201%20in%203%20adults,not%20result%20in%20serious%20injury.

3. Varela-Vasquez LA, Minobes-Molina E, Jerez-Roig J. Dual-task exercises in older adults: A structured review of current literature. J Frailty Sarcopenia Falls. 2020;5(2):31–7.

4. Falls: applying All Our Health 2022 15/07/2024. Available from: https://www.gov.uk/government/publications/falls-applying-all-our-health/falls-applying-all-our-health.

5. Chantanachai T, Sturnieks DL, Lord SR, Payne N, Webster L, Taylor ME. Risk factors for falls in older people with cognitive impairment living in the community: Systematic review and meta-analysis. Ageing Res Rev. 2021;71:101452.

6. Allali G, Launay CP, Blumen HM, Callisaya ML, De Cock AM, Kressig RW, et al. Falls, Cognitive Impairment, and Gait Performance: Results From the GOOD Initiative. J Am Med Dir Assoc. 2017;18(4):335–40.

7. Karen Z. H. Li, Halina Bruce, Downey R. Cognition and Mobility With Aging. Oxford Research Encyclopedia of Psychology. 2018.

8. Buchman AS, Boyle PA, Leurgans SE, Barnes LL, Bennett DA. Cognitive Function Is Associated With the Development of Mobility Impairments in Community-Dwelling Elders. The American Journal of Geriatric Psychiatry. 2011;19(6):571–80.

9. Demnitz N, Esser P, Dawes H, Valkanova V, Johansen-Berg H, Ebmeier KP, Sexton C. A systematic review and meta-analysis of cross-sectional studies examining the relationship between mobility and cognition in healthy older adults. Gait Posture. 2016;50:164–74.

10. Buchman AS, Boyle PA, Leurgans SE, Barnes LL, Bennett DA. Cognitive function is associated with the development of mobility impairments in community-dwelling elders. Am J Geriatr Psychiatry. 2011;19(6):571–80.

11. Demnitz N, Hogan DB, Dawes H, Johansen-Berg H, Ebmeier KP, Poulin MJ, Sexton CE. Cognition and mobility show a global association in middle- and late-adulthood: Analyses from the Canadian Longitudinal Study on Aging. Gait Posture. 2018;64:238–43.

12. Muir-Hunter SW, Wittwer JE. Dual-task testing to predict falls in community-dwelling older adults: a systematic review. Physiotherapy. 2016;102(1):29–40.

13. Bayot M, Dujardin K, Dissaux L, Tard C, Defebvre L, Bonnet CT, et al. Can dual-task paradigms predict Falls better than single task? - A systematic literature review. Neurophysiol Clin. 2020;50(6):401–40.

14. Wang X, Pi Y, Chen P, Liu Y, Wang R, Chan C. Cognitive motor interference for preventing falls in older adults: a systematic review and meta-analysis of randomised controlled trials. Age Ageing. 2015;44(2):205–12.

15. Sullivan AN, Lachman ME. Behavior Change with Fitness Technology in Sedentary Adults: A Review of the Evidence for Increasing Physical Activity. Front Public Health. 2016;4:289.

16. Callisaya ML, Jayakody O, Vaidya A, Srikanth V, Farrow M, Delbaere K. A novel cognitive-motor exercise program delivered via a tablet to improve mobility in older people with cognitive impairment - StandingTall Cognition and Mobility. Exp Gerontol. 2021;152:111434.

17. Papi E, Chiou SY, McGregor AH. Feasibility and acceptability study on the use of a smartphone application to facilitate balance training in the ageing population. BMJ Open. 2020;10(12):e039054.

18. Valenzuela T, Okubo Y, Woodbury A, Lord SR, Delbaere K. Adherence to Technology-Based Exercise Programs in Older Adults: A Systematic Review. J Geriatr Phys Ther. 2018;41(1):49–61.

19. Netz Y, Yekutieli Z, Arnon M, Argov E, Tchelet K, Benmoha E, Jacobs JM. Personalized Exercise Programs Based upon Remote Assessment of Motor Fitness: A Pilot Study among Healthy People Aged 65 Years and Older. Gerontology. 2022;68(4):465–79.

20. Smith-Ray RL, Makowski-Woidan B, Hughes SL. A randomized trial to measure the impact of a community-based cognitive training intervention on balance and gait in cognitively intact Black older adults. Health Educ Behav. 2014;41(1 Suppl):62S–9S.

21. Rasche P, Wille M, Brohl C, Theis S, Schafer K, Knobe M, Mertens A. Prevalence of Health App Use Among Older Adults in Germany: National Survey. JMIR Mhealth Uhealth. 2018;6(1):e26.

22. McGarrigle L, Boulton E, Todd C. Map the apps: a rapid review of digital approaches to support the engagement of older adults in strength and balance exercises. BMC Geriatr. 2020;20(1):483.

23. Wilson J, Heinsch M, Betts D, Booth D, Kay-Lambkin F. Barriers and facilitators to the use of e-health by older adults: a scoping review. BMC Public Health. 2021;21(1):1556.

24. Hughes KJ, Salmon N, Galvin R, Casey B, Clifford AM. Interventions to improve adherence to exercise therapy for falls prevention in community-dwelling older adults: systematic review and meta-analysis. Age Ageing. 2019;48(2):185–95.

25. Wang RY, Huang YC, Zhou JH, Cheng SJ, Yang YR. Effects of Exergame-Based Dual-Task Training on Executive Function and Dual-Task Performance in Community-Dwelling Older People: A Randomized-Controlled Trial. Games Health J. 2021;10(5):347–54.

26. PEAK-Brain Training App. https://www.peak.net/science/.

27. Ltd. SL. Peak (Version 5.34.9) [Mobile application software]. 2024.

28. Sim J, Lewis M. The size of a pilot study for a clinical trial should be calculated in relation to considerations of precision and efficiency. J Clin Epidemiol. 2012;65(3):301–8.

29. Teare MD, Dimairo M, Shephard N, Hayman A, Whitehead A, Walters SJ. Sample size requirements to estimate key design parameters from external pilot randomised controlled trials: a simulation study. Trials. 2014;15:264.

30. Lancaster GA, Dodd S, Williamson PR. Design and analysis of pilot studies: recommendations for good practice. J Eval Clin Pract. 2004;10(2):307–12.

31. Alexander NB, Hausdorff JM. Guest editorial: linking thinking, walking, and falling. J Gerontol A Biol Sci Med Sci. 2008;63(12):1325–8.

32. Liu Y, Chan JS, Yan JH. Neuropsychological mechanisms of falls in older adults. Front Aging Neurosci. 2014;6:64.

33. Clouston SA, Brewster P, Kuh D, Richards M, Cooper R, Hardy R, et al. The dynamic relationship between physical function and cognition in longitudinal aging cohorts. Epidemiol Rev. 2013;35(1):33–50.

34. Divandari N, Bird ML, Vakili M, Jaberzadeh S. The Association Between Cognitive Domains and Postural Balance among Healthy Older Adults: A Systematic Review of Literature and Meta-Analysis. Curr Neurol Neurosci Rep. 2023;23(11):681–93.

35. Hawley-Hague H, Horne M, Campbell M, Demack S, Skelton DA, Todd C. Multiple levels of influence on older adults’ attendance and adherence to community exercise classes. Gerontologist. 2014;54(4):599–610.

36. Lucidi F, Grano C, Barbaranelli C, Violani C. Social-cognitive determinants of physical activity attendance in older adults. J Aging Phys Act. 2006;14(3):344–59.

37. Avan BI, Berhanu D, Umar N, Wickremasinghe D, Schellenberg J. District decision-making for health in low-income settings: a feasibility study of a data-informed platform for health in India, Nigeria and Ethiopia. Health Policy Plan. 2016;31 Suppl 2(Suppl 2):ii3–ii11.

38. Shumway-Cook A, Brauer S, Woollacott M. Predicting the probability for falls in community-dwelling older adults using the Timed Up & Go Test. Physical Therapy. 2000;80(9):896–903.

39. Zasadzka E, Borowicz AM, Roszak M, Pawlaczyk M. Assessment of the risk of falling with the use of timed up and go test in the elderly with lower extremity osteoarthritis. Clin Interv Aging. 2015;10:1289–98.

40. Tomaszewski Farias S, Mungas D, Harvey DJ, Simmons A, Reed BR, Decarli C. The measurement of everyday cognition: development and validation of a short form of the Everyday Cognition scales. Alzheimers Dement. 2011;7(6):593–601.

41. Farias ST, Weakley A, Harvey D, Chandler J, Huss O, Mungas D. The Measurement of Everyday Cognition (ECog): Revisions and Updates. Alzheimer Dis Assoc Disord. 2021;35(3):258–64.

42. Delbaere K, Close JC, Mikolaizak AS, Sachdev PS, Brodaty H, Lord SR. The Falls Efficacy Scale International (FES-I). A comprehensive longitudinal validation study. Age Ageing. 2010;39(2):210–6.

43. Herdman M, Gudex C, Lloyd A, Janssen M, Kind P, Parkin D, et al. Development and preliminary testing of the new five-level version of EQ-5D (EQ-5D-5L). Qual Life Res. 2011;20(10):1727–36.

44. Garfield K, Thorn JC, Noble S, Husbands S, Hollingworth W. Development of a brief, generic, modular resource-use measure (ModRUM): piloting with patients. BMC Health Serv Res. 2023;23(1):994.

45. Goodyear VA, Boardley I, Chiou SY, Fenton SAM, Makopoulou K, Stathi A, et al. Social media use informing behaviours related to physical activity, diet and quality of life during COVID-19: a mixed methods study. BMC Public Health. 2021;21(1):1333.

46. Stathi A, Greaves CJ, Thompson JL, Withall J, Ladlow P, Taylor G, et al. Effect of a physical activity and behaviour maintenance programme on functional mobility decline in older adults: the REACT (Retirement in Action) randomised controlled trial. Lancet Public Health. 2022;7(4):e316–e26.

47. IBM SPSS Statistics for Windows. 29.0.2.0 ed. Armonk, NY: IBM Corp 2023.

48. Tinetti ME, Baker DI, McAvay G, Claus EB, Garrett P, Gottschalk M, et al. A multifactorial intervention to reduce the risk of falling among elderly people living in the community. N Engl J Med. 1994;331(13):821–7.

49. Anne Shumway-Cook MW. Translating research into clinical practice. Abstracts of the Seventh International Symposium on Osteoporosis. April 18-22, 2007. Washington, DC, USA. Osteoporos Int. 2007;18 Suppl 2:S193–244.

50. Percentage of homes and individuals with technological equipment 2022 19/08/2024. Available from: https://www.beta.ons.gov.uk/aboutus/transparencyandgovernance/freedomofinformationfoi/percentageofhomesandindividualswithtechnologicalequipment.

